# Feasibility and efficacy of personalized cardiorespiratory and strength exercise in Duchenne muscular dystrophy: a pilot study

**DOI:** 10.64898/2026.09.08.26361717

**Authors:** Meghana Bomma, Donovan Lott, Sean Forbes, Victoria Del Toro, Diego Maldonado-Puebla, Mallory Paul, Ruby Sullivan, Andrew Carvill, Aiden Foy, Renata Shih, John Anthony Coppola, Carmen Leon-Astudillo, Glenn Walter, Michael Daniels, Kimberly Stubbs, Warren Dixon, H. Lee Sweeney, Tanja Taivassalo

**Author notes:** **Correspondence address:** Tanja Taivassalo, Clinical Translational Research Building, Room 2214, 2004 Mowry Road, University of Florida, Gainesville, Florida 32610.

## Abstract

**Background:** Duchenne muscular dystrophy (DMD) results from absence of dystrophin, causing sarcolemmal instability, progressive muscle and mitochondrial dysfunction and reduced cardiorespiratory fitness and bone density. Exercise may target these multisystem abnormalities but remains underutilized due to historical safety concerns and lack of disease-specific guidelines for optimal dosing. We conducted an exploratory trial using personalized exercise prescription to assess adherence, safety and preliminary efficacy in DMD.

**Methods:** Six ambulatory boys underwent 6-months of home-based, remotely-supervised leg cycling and isometric strengthening using individualized frequency, intensity and time parameters. Cycling was set to 40-60% of heart rate and strengthening to 50% maximal voluntary contraction (MVC). Safety was assessed by creatine kinase (CK), muscle MRS T_2_, and adverse events. Primary outcomes were 6-month change in peak oxygen uptake (VO₂) and submaximal-cycling HR; secondary outcomes included muscle fat fraction, cross-sectional area (CSA) and peak strength; exploratory outcomes included bone density, pulmonary function and quality-of-life. Peak VO_2_ and CSA were contextualized against untreated DMD controls.

**Results:** All participants completed the intervention with high adherence (88.5%) and no adverse events. Cycling averaged 2-3 times/week at HR 148±8 bpm for 21.9±3.9 minutes; strengthening averaged 1.8±0.6 sessions/week at 50% MVC. CK and MRS T_2_ remained stable. Peak VO₂ and submaximal-cycling HR improved, CSA increased relative to controls, and hamstring strength increased. Bone density, forced vital capacity and quality-of-life scores showed favorable trends.

**Discussion:** Appropriately dosed exercise is feasible, safe and may produce multisystem physiological benefits in boys with DMD, supporting personalized, physiologically monitored training as a complementary therapeutic strategy.

Trial registry name and number: Exercise as Therapy for DMD; NCT04322357

## INTRODUCTION

Duchenne muscular dystrophy (DMD) is a severe, rapidly progressive X-linked neuromuscular disease with a male newborn incidence of 1 in 20,000^1^. It is caused by gene mutations leading to absence of functional dystrophin in skeletal and cardiac muscle^2^. This loss results in mechanical instability, increased membrane damage, mitochondrial dysfunction and repeated cycles of myofiber degeneration and regeneration that drive progressive replacement of muscle with fat and fibrotic tissue ^3, 4^. Clinically, DMD is characterized by severe muscle weakness and fatigability, loss of ambulation in teenage years and death from respiratory or cardiac failure in subsequent decades^5, 6^. Beyond skeletal muscle involvement, DMD is associated with profoundly reduced cardiorespiratory fitness^7^, diminished bone density with elevated fracture risk^8^, and high prevalence of metabolic dysfunction^9,10^. These deficits are compounded by physical inactivity and long-term corticosteroid use (current standard of care) further worsening clinical outcomes and quality of life^11–14^. While emerging gene therapies offer partial restoration though microdystrophin, they yield variable effects and do not target the multi-systemic deficits highlighting the need for adjuvant therapies targeting the broader presentation of DMD pathophysiology.

Exercise represents a potentially valuable yet underutilized therapeutic strategy in DMD with capacity to impact skeletal muscle, cardiorespiratory, and bone health. Historical concerns regarding contraction-induced injury and overuse weakness^15^ constrained earlier investigations^25–27^. More recent studies using submaximal exercise paradigms suggest that it may preserve function in boys with DMD without evidence of overuse or injury^16–21^. Nevertheless, the therapeutic efficacy of exercise, particularly with respect to improvements in muscle strength or endurance, remains uncertain^22^. This uncertainty reflects the limited number of studies and small sample sizes, limited use of quantitative outcomes assessing the mechanistic impact and the absence of defined exercise guidelines tailored specifically for DMD. Precise definition of exercise dose is essential to balance the susceptibility to dystrophin-deficient muscle to damage with the need to activate adaptive signaling pathways that may confer benefit^23^. This necessitates a personalized approach, analogous to precision medicine^24^, in which exercise prescription is individualized to physiological capacity and disease-specific vulnerability. The FITT principal (Frequency, Intensity, Time, Type)^25^ is widely applied to optimize exercise prescription across clinical populations,^26^ and international consensus FITT-based recommendations have recently been defined for cardiorespiratory and resistance exercise in adults with muscle diseases^27^. Comparable guidance is currently lacking for DMD, representing a critical gap in defining safe and effective exercise interventions for this population.

Our group has begun to examine the feasibility of precision-based exercise dosing in DMD. We previously demonstrated the safety and preliminary efficacy of individualized isometric strength training using quantitative outcome measures sensitive to muscle pathophysiology in a small cohort of DMD^20^. More recently, we established feasibility of prescribing cardiorespiratory exercise using personalized heart rate (HR) zones to guide submaximal cycling intensity^28^. Building on this work, the present study reports results of an exploratory intervention combining cardiorespiratory and strength training using a fully personalized exercise prescription in ambulatory boys with DMD. The objectives were to 1) assess feasibility of personalizing exercise using FITT factors; 2) evaluate safety and obtain preliminary efficacy data on muscle pathophysiology and cardiorespiratory fitness; and 3) explore potential effects on pulmonary function, bone density and quality of life. We hypothesized that 6-months of personalized exercise training would result in improvements in cardiorespiratory fitness and preservation of muscle health, reflected b stabilization of disease-related outcomes.

## METHODS

### Study Design and Approvals

This single-site (University of Florida, UF) exploratory longitudinal cohort study of home-based exercise in boys with DMD included visits for outcome assessment at baseline, 3 and 6 months after starting the intervention. To account for expected disease progression over this time frame, independent untrained (UT) DMD control cohorts were evaluated for selected primary and secondary outcomes. Outcome analyses were performed blinded to the timepoint. This study was registered at ClinicalTrials.gov (NCT04322357) and approved by the UF Institutional Review Board (Project: 201901339). Untrained DMD controls for muscle cross-sectional area (control cohort 1) were enrolled in a separate study arm and UT DMD controls for cardiorespiratory variables (control cohort 2) were enrolled in a separate study (Project number: 202101689). Written informed consent and assent were obtained for each participant. All procedures conformed to the Declaration of Helsinki.

### Participants

Participants were recruited through the Center for Neuromuscular and Rare Diseases at UF and ClinicalTrials.gov. Inclusion criteria were genetically confirmed DMD, aged 6 to 9 years, ambulatory (≥ 100 meters independently) and on a stable glucocorticoid regimen (a minimum of 6 months)^14^. Exclusion criteria included unstable medical conditions contraindicating exercise as indicated by the American College of Sports Medicine (ACSM) guidelines^29^, significant cardiomyopathy or conduction abnormalities, prior rhabdomyolysis, MRI contraindications, investigational drug or gene-therapy participation and use of growth hormone, carnitine, creatine or other performance-enhancing agents. Identical criteria were applied to UT control cohorts, who did not undergo structured exercise training.

### Intervention

The home-based intervention was remotely supervised and included two modalities (leg cycling and isometric strengthening) involving predominantly the quadriceps and hamstring muscles (Figure 1). Cycle exercise was based on FITT parameters recommended by ACSM for deconditioned and heart failure populations^25, 30^. Phase 1 of the intervention (Months 0-3) involved active cycling, 3 sessions per week at 40-50% heart rate reserve (HRR), 15-30 minutes intermittently or continuously. HRR (peak–resting HR) was calculated using measured peak HR from baseline cardiopulmonary exercise testing (CPET)^30^. The Karvonen formula was used to establish the personalized target HR zone (40-60% HRR+HR rest). Phase 2 (Months 3-6) involved a combination of modalities where cycling was reduced to 1-2 sessions per week (intensity between 50-60% HRR) and isometric knee extension (KE) and knee flexion (KF) strengthening was added using FITT parameters similar to our previous trial ^20^: 1-2 sessions per week at 50% MVC, 4 sets of 6 repetitions on each leg at 60 degrees of KF. Between sets, a rest of at least 1-minute was provided.

**Figure 1.**
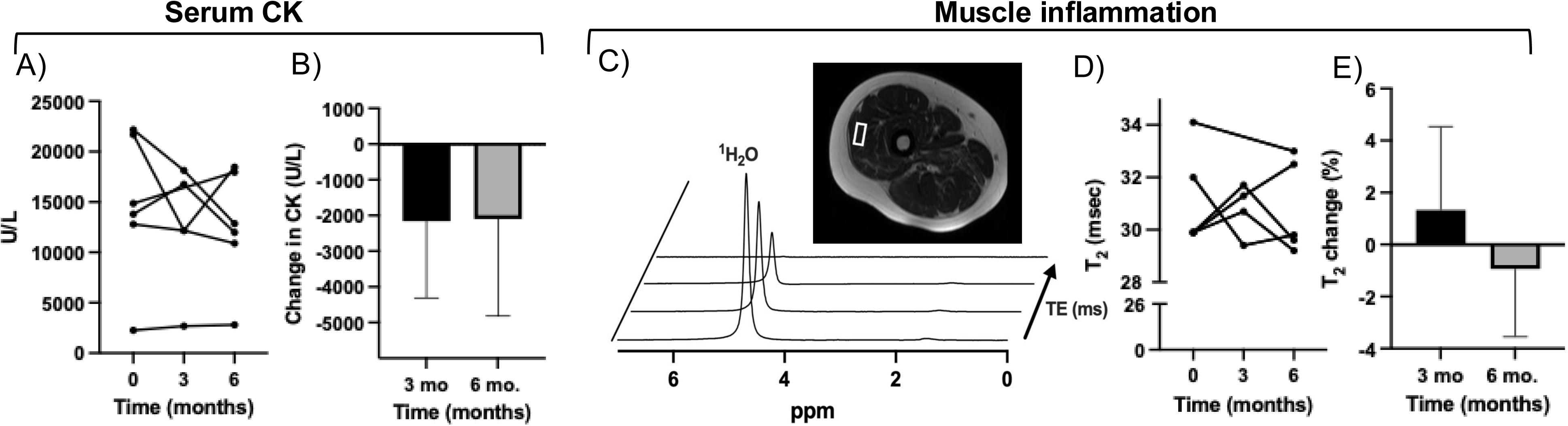
Study design and exercise intervention. A) Overview of the home-based, remotely supervised 6-month exercise intervention using personalized FITT parameters (Frequency, Intensity, Time, Type). Outcome measures assessed at baseline (time 0), 3 and 6-months are indicated. *Figure created in* https://BioRender.com Phase 1 consisted of active cycling performed on a customized motor-assisted ergometer (B) in five participants, with the level of assistance adjusted to maintain the target HR zone^28^. Due to mechanical issues, one participant used a non-motorized tricycle (Rifton Equipment, New York, USA) (C) with cycling intensity adjusted using gears to maintain the target HR zone. Phase 2 consisted of active cycling in combination with (D) isometric knee extension and flexion strengthening performed using a customized leg extension/flexion device as previously described^20^. Abbreviations: VO_2_=rate of oxygen utilization; HR=heart rate; CK=creatine kinase; MRS-T_2_=magnetic resonance spectroscopy transverse relaxation time constant; FF=fat fraction; CSA=cross-sectional area; KE=knee extension; KF=knee flexion; PFT=pulmonary function testing; DEXA= Dual-energy x-ray absorptiometry; PCrτ= phosphocreatine time constant (tau) of phosphocreatine recovery; PROM=patient reported outcome measures; HRR=heart rate reserve min=minutes; MVC=maximal voluntary contraction, min = minutes.

Participants were instructed to exercise at home on alternate days within a target perceived exertion ranging from 3-7 (Borg 1-10 scale). Cycling was conducted on a custom-built, motor-assisted ergometer with real-time remote adjustment of assistance to maintain target HR as previously described^28^. Strength training used a leg extension/flexion device customized for isometric movements in children^20^. Participants were provided with the cycle and strengthening devices, a laptop computer (for remote data collection and live video-streaming using Zoom video [Zoom communications Inc, Denver, USA]^31^) and HR chest strap and watch (Polar H9 and Unite; Polar Electro, Finland). For every exercise session, the study team communicated with the participant in real-time using Zoom video to monitor symptoms (muscle pain or soreness, dyspnea and/or overall fatigue), ratings of perceived exertion and HR, adjust motor assistance based on HR (cycling) and target MVC (strengthening) and motivate the participant. Data files were transferred to a secure drive at UF for subsequent analysis of exercise metrics and adherence.

### Outcomes Measures

Safety and efficacy outcomes were collected at baseline, 3 and/or 6-months and adverse events (i.e. any undesirable occurrence relating to the intervention or study procedures) were monitored throughout the study. At baseline, demographic information including past and current medications was collected and participants underwent an echocardiogram.

To monitor safety, resting levels of serum creatine kinase (CK) and the magnetic resonance spectroscopy (MRS)-transverse relaxation time constant (MRS-T_2_, a marker of muscle damage, inflammation and edema^20, 32–34^) in the vastus lateralis (VL) were measured at baseline, 3 and 6-months. Prespecified thresholds indicative of muscle injury included elevation in CK of 7,000 U/L above baseline and ≥20% increase in MRS-T_2_ (similar to our previous strength trial^20^).

Preliminary efficacy was determined by the 6-month change in co-primary outcomes peak VO_2_ and submaximal exercise HR; both established markers of cardiorespiratory training response^25^. To quantify peak VO_2_, cardiopulmonary exercise testing (CPET) was performed using ACSM guidelines at baseline and 6-months where participants underwent symptom-limited incremental exercise to maximal effort on a recumbent stationary electronically braked cycle ergometer (Lode Corrival, Lode BV, Gronigen, The Netherlands). Continuous 12-lead ECG (CardioSuite Cardio Card, Nasiff Associates, Inc, New York, USA) and breath-by-breath gas exchange (ParvoMedics, TrueOne, Salt Lake City, Utah, USA) were collected using a CPET protocol recently shown to be valid and reproducible in DMD by our group^7^. Peak VO_2_ and watts (W) were defined as the highest 30-second average achieved during CPET. Resting and peak HR attained at baseline were used to quantify HRR and prescribe cycle training intensity. DMD control cohort 2 underwent the same peak-exercise protocol. In a separate test at each visit, submaximal constant load cycling was performed at ∼ 30-50% peak workload. Average HR and time within target HR zone were recorded. At 3 and 6-months, the constant-load test was repeated at the same workload and pedaling speed. Reduction in average HR and increase in interval time after training were considered outcomes of improved submaximal exercise tolerance.

Secondary outcomes included the 6-month change in markers of skeletal muscle pathology: fat fraction (FF) and cross-sectional area (CSA) of the quadriceps and hamstrings (recruited during prescribed exercises) and KE and KF strength. For the former, magnetic resonance imaging (MRI) was performed at 3T (MR7700, Philips, Best, The Netherlands) using multi-slice 3-point chemical shift encoded (Dixon) imaging to map both fat and water signals protocols as previously established by our group^34,35, 36^. Briefly, Dixon sequences were run using a fast field echo (FFE) pulse sequence with a 20 flip angle at 3 different echo times (TR/TE = 430/8.06 ms, 9.21 ms, 10.36 ms) over ∼30 axial slices (4-mm slice thickness, 1 mm gap) using mDixon (Philips) and FOV adjusted depending on subject anatomy. Water and fat maps were reconstructed using the standard Philips in-line processing (mDixon) with the fat map derived using a 7 peak fat model ^37^. The MR images were analyzed using Horos software v4.0.0 (horosproject.org, Annapolis, MD) to calculate cross-sectional area (CSA) and fat fraction (FF) of three slices chosen in the mid-thigh region identified by the landmark in which the biceps femoris short head muscle emerges^38^. We also calculated contractile CSA as [CSA x [1-FF]) to reflect the amount of functional muscle tissue by accounting for infiltrated fat.^39^ ^1^H-MRS ^1^H_2_O T_2_ data were acquired using stimulated-echo acquisition mode (STEAM) from the vastus lateralis (4 repeated TE’s nonlinearly spaced from 11–243 ms; TR = 9 s; NA = 4); voxel size was optimized for each individual’s muscle size and ^1^H_2_O peak was fit with monoexpontial model ^40^. UT controls underwent an identical MRI protocol to quantify CSA. For strength, isometric maximal voluntary torque was quantified on a fixed dynamometer (Biodex Medical Systems, Shirley, NY) for each leg as previously described^20^. The highest torque value of five trials per leg was used with data representing the average of left and right leg.

Exploratory outcomes included pulmonary function, lumbar spine bone density, ambulatory motor function, the time constant of phosphocreatine recovery and quality of life. Forced vital capacity (FVC) and forced expiratory volume in 1 second (FEV_1_) (Carefusion Microlab Portable spirometer, Micro Medical Limited, Kent, United Kingdom) as well as maximal inspiratory pressure (MIP) and maximal expiratory pressure (MEP) (Carefusion MicroRPM, Kent, United Kingdom) were measured according to American Thoracic Society (ATS) standards^41,42^. Dual-energy x-ray absorptiometry (DEXA, Lunar Prodigy Advance, GE Healthcare) was used to quantify lumbar spine bone density Z-score as well as total leg lean mass and fat percentage. The 10m walk/run time, North Star Ambulatory Assessment [NSAA] and 6-minute walk distance [6MWD]) were used to assess motor function assessed using validated standardized instructions and methodology^43–45^. ^31^P-MRS was applied to measure the phosphocreatine recovery time constant (PCrτ), a metric used to monitor mitochondrial ATP production and reflect muscle oxidative capacity ^46, 47^ within the VL. For these acquisitions, each participant was positioned supine with their right knee flexed at ∼60° and ankle linked to strap with force transducer for isometric leg extension contractions. A 14 cm flex ^31^P-MRS surface coil was placed on the quadriceps region of the upper leg. An unlocalized ^31^P-MRS sequence with the pulse excitation centered between PCr and γATP (adiabatic pulse, bandwidth: 2500 Hz, 2048 points, TR: 3 s, NSA: 4) was obtained. Dynamic unlocalized ^31^P-MRS spectra were acquired for ten minutes continuously. During this scan, the isometric exercise protocol was performed, and included 2 min of rest, 1 min of repeated maximal isometric contractions (0.5 Hz) followed by 7 min of recovery. ^31^P-MRS data were analyzed using the AMARES quantitation algorithm in jMRUi (v. 7.0)^48, 49^. Estimated starting values and prior knowledge were utilized for PME, Pi, PDE, PCr, and γ-, α-, and β-ATP. Intracellular pHi was calculated by the chemical shift between Pi relative to PCr^50^. A mono-exponential model was used to PCrτ.

Finally, quality of life was assessed using the Pediatric Quality of Life Inventory^TM^ 3.0 (PedsQL-DMD) parent module and Multidimensional Fatigue Scale (PedsQL-MDF) patient module, where a higher score indicates better QoL or less impact of fatigue on QoL. To complement these validated scales, a caregiver-reported 7-point Likert Impression of Change scale evaluated perceived changes energy level, strength, endurance, play and mood.

### Statistical analysis

Non-parametric analyses were conducted using Graphpad prism version 10 (Boston, Massachusetts, USA). Differences in changes in safety markers (baseline, 3-months and 6-months) were tested using the Wilcoxon signed rank test with Bonferroni correction (for the two tests). The 6-month change in primary outcome (Δ in peak VO_2_) and all secondary and exploratory outcomes were also tested using the Wilcoxon signed rank test. To compare the trajectories of submaximal HR (co-primary outcome) during constant-load cycling (Fig 2E), we fit mixed models with a random intercept and a quadratic trajectory; we tested for differences in baseline, 3-months and 6-months using likelihood ratio tests. To test whether the 6-month change in peak VO_2_ and muscle CSA differed between the trained and UT DMD cohorts, a Mann-Whitney test was conducted.

**Figure 2:**
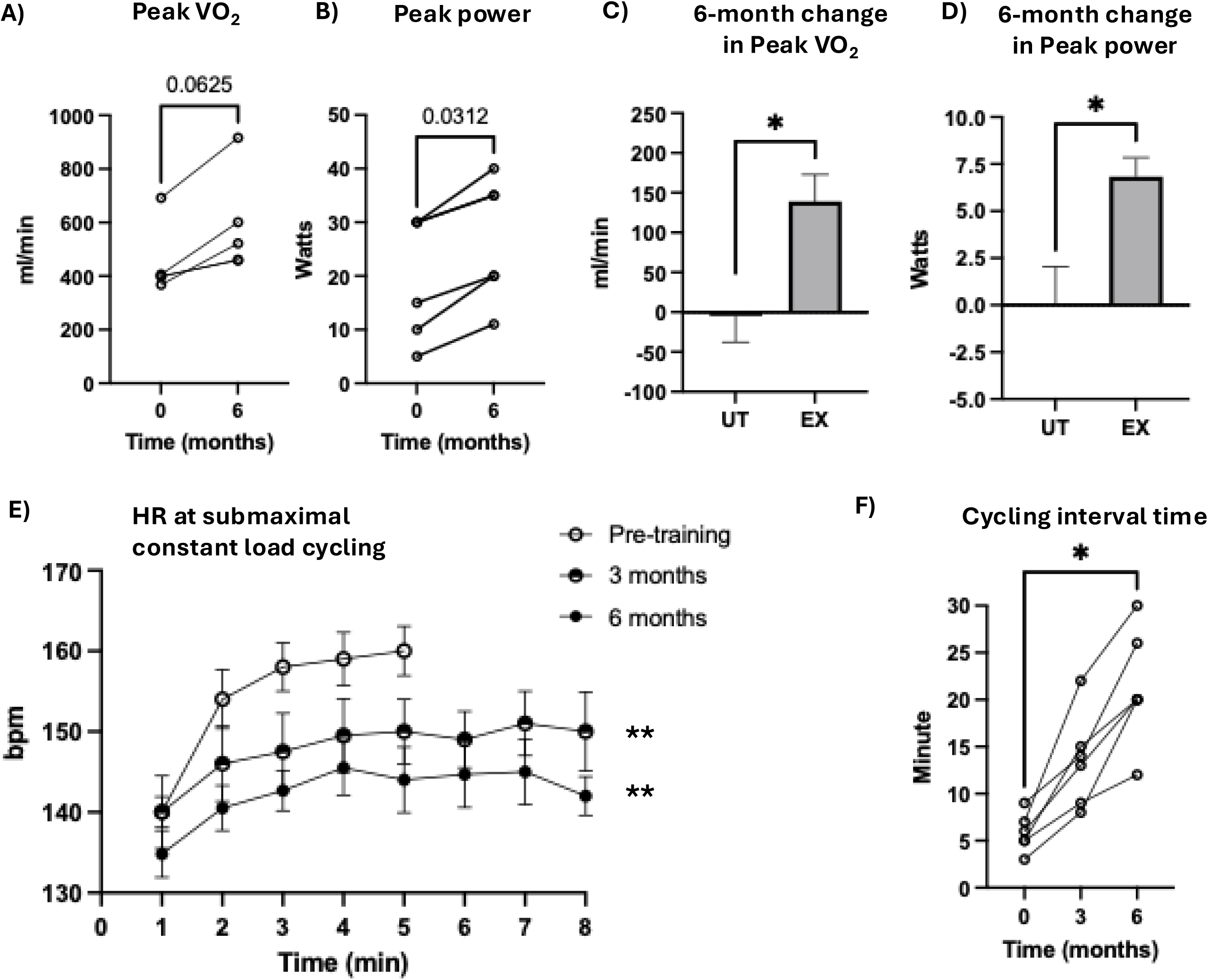
Safety makers of muscle injury following 6-months of personalized exercise in DMD. A) Resting serum CK levels shown per individual participants at baseline and after training. B) Mean change in CK (± SEM) at 3 and 6 months relative to baseline. C) Representative MRS ^1^H_2_O T_2_ spectra acquired from a voxel positioned in the vastus lateralis. D) MRS ^1^H_2_O T_2_ values shown for individual participant at baseline and following training. E) Mean MRS-T_2_ values (± SEM) at baseline, 3 and 6 months post intervention.

## RESULTS

### Participants

Six participants in the intervention arm completed the exercise program with no adverse events (demographics shown in Table 1). Five participants completed all 3 study visits; one participant (EX01) was unable to travel for the 3-month visit. There was a 4-month delay in the initiation of training for one participant (EX-04) due to equipment shipment-related issues. The first 5 enrolled participants completed the cycling protocol using the custom-built motor-assisted ergometer. During the latter two months of training, two participants (EX-04 and EX-05) experienced mechanical issues with the ergometer, necessitating an increased volume of strength training to maintain a comparable overall exercise volume relative to other participants (see Supplemental table). One participant (EX-06) experienced mechanical issues after 1 month of training and subsequently transitioned to a commercially available, non-motor assisted adaptive tricycle (Rifton Equipment, New York, USA) for the remainder of the intervention. This resulted in a 1-week interruption in training. The use of active cycling was supported by our prior work demonstrating feasibility of active cycling in boys with DMD^28^. For the non-exercised DMD controls, cohort 1 included 7 boys (6.1±1.2 years) and cohort 2 included 4 boys (11.8±3.5 years). All control participants were receiving a stable corticosteroid regimen and were not receiving other FDA approved or investigational therapies.

**Table 1.** Baseline demographics of boys with DMD who underwent the 6-month exercise intervention and untrained DMD controls.

| **DMD-EX** | **Age range, Years** | **BMI, Kg/m<sup>2</sup>** | **Medications** | **EF, %** | **6MW D, meters** | **10 m time, sec** | **NSAA score** |
| --- | --- | --- | --- | --- | --- | --- | --- |
| EX01 | 5-9 | 22.5 | Daily prednisone | 68 | 387 | 4.06 | 28 |
| EX02 | 5-9 | 17.1 | Daily deflazacort, trazodone | 69 | 425 | 3.31 | 29 |
| EX03 | 5-9 | 17.9 | Daily prednisone | 70 | 509 | 2.15 | 27 |
| EX04 | 5-9 | 21.5 | Twice-weekly prednisone, lisinopril, carvedilol | 50 | 302 | 5.25 | 25 |
| EX05 | 5-9 | 17.2 | Twice-weekly prednisone | 67 | 366 | 4.0 | 19 |
| EX06 | 5-9 | 26.5 | Daily deflazacort, viltolarsen | 66 | 177 | 11.5 | 6 |
| **AVG** | 5-9 | **20.5** |  | **65** | **361** | **5.0** | **22.3** |
| **stdev** |  | **3.7** |  | **7.5** | **113** | **3.3** | **8.8** |
| UT01 | 5-9 | 14.9 | Twice-weekly prednisone | NA | 405 | 3.29 | 26 |
| UT02 | 5-9 | 16.0 | Twice-weekly prednisone | NA | NA | 3.94 | NA |
| UT03 | 5-9 | 17.5 | Twice-weekly prednisone | NA | 264 | 5.97 | 18 |
| UT04 | 5-9 | 17.1 | Twice-weekly prednisone | NA | 480 | 3.96 | 25 |
| UT05 | 5-9 | 15.2 | Twice-weekly prednisone | NA | NA | 5.55 | 18 |
| UT06 | 5-9 | 17.9 | Twice-weekly deflazacort | NA | 350 | 5.65 | 20 |
| UT07 | 5-9 | 17.1 | Twice-weekly prednisone | NA | NA | 5.56 | 18 |
| **AVG** | 5-9 | **16.5** |  |  | **375** | **4.8** | **20.8** |
| **stdev** |  | **1.2** |  |  | **91** | **1.1** | **3.7** |
| UT08 | 5-9 | 19.6 | Twice-weekly prednisone | 64.5 | 392 | 4.34 | 26 |
| UT09 | 10-14 | 15.4 | Daily prednisone, lisinopril | 66.2 | 320 | 7.7 | 21 |
| UT10 | 15-19 | 19.8 | Daily prednisone, lisinopril, eplerenone | 74.2 | 444 | 3.8 | 30 |
| UT11 | 10-14 | 16.4 | Deflazacort, lisinopril, eplerenone, givinostat | 72 | 401 | 5.0 | NA |
| **AVG** | 10-14 | 17.8 |  |  | 389.3 | 5.2 | 25.7 |
| **stdev** |  | 2.2 |  |  | 51.4 | 1.7 | 4.5 |

BMI=body mass index; Kg=kilograms; m<sup>2</sup>= meters squared; EF = ejection fraction; 6MWD=six minute walk distance; 10 m = 10 meter; sec=seconds; NSAA=North Star Ambulatory Assessment; AVG=average; stdev=standard deviation; UT=untrained; NA=not available.

### Feasibility of Intervention

Details on individual participant adherence and personalized exercise parameters for cycling and isometric strengthening are found in the Supplemental Table.

#### Frequency

Participants completed a mean of 69.0±11.3 of 78 prescribed exercise sessions over the 6-month intervention, corresponding to an overall adherence of 88.5% and consistent with the prescribed frequency of approximately 3 sessions a week. This included 47±15 cycling sessions and 22±7.8 strength training sessions. Adherence to cycling sessions during Phase 2 showed greater variability due to mechanical issues with exercise equipment. As a result, two participants (EX-4 and EX-5) completed a higher number of strength sessions (2 to 3 per week) relative to cycling compared to cycling (once per week).

#### Intensity

The target cycling HR zone was calculated as 137-151 bpm based on a mean participant age of 7.9±1.1 years, resting HR of 111.0±7.0 bpm and peak measured HR of 177.0±11.0 bpm. Participants trained at a mean cycling HR of 148±8 bpm, corresponding to 57.5±11.7% HRR, indicating feasibility of prescribing exercise intensity within a 40-60% HRR range. The mean power output achieved at this HR intensity was 5.1±1.0 watts. Strength training intensity was prescribed at 50% maximal voluntary contraction and corresponded to mean torques of 11.2±7.1 Nm for KE and 12.6±8.5 Nm respectively for KF.

#### Time

Cycling exercise was prescribed for 15-30 minutes per session within the target HR zone. During the initial phase of training, cycling was performed in shorter intervals interspersed with rest periods. These rest breaks of 1-5 minutes were taken when HR exceeded the prescribed upper limit. In the first month of training, mean interval duration in the HR zone was 5.2±1.5 minutes, with a total cycling duration of 15.6±2.5 minutes per session. Exercise time progressed over the intervention, such that by month 6, the mean continuous time within the HR zone increased to 20.4±6.4 minutes and total session duration to 28.6±1.9 minutes. The mean distance cycled over the 6-month intervention was 1.3±0.2 miles. Strength training was prescribed as 4 sets of 6 repetitions for both KE and KF on each leg, with a minimum of 1-minute rest between sets, resulting in 96 repetitions per training session. Over the 6-month training period, participants completed a mean total of 1,655±392 isometric leg repetitions.

### Safety and Muscle Injury Markers

Baseline resting serum CK and VL MRS T_2_ were 14,592±7264 U/L and 31.2±1.9 msec respectively. Following the intervention, no participant met predefined thresholds indicative of muscle injury. Mean changes in CK were -2210±4818 U/L at 3-months and -2099±6668 U/L at 6-months. Corresponding changes in VL MRS T_2_ were 1.3±6.4% at 3-months and -0.9±5.8% at 6-months (Figure 2). Cardiac ejection fraction (baseline: 65.6±8.8%; 6-months: 65.0±12.3%) and BMI (baseline: 20.4±3.7 kg/m^2^; 6-months: 21.8±3.9 kg/m^2^) were unchanged.

### Cardiorespiratory fitness

Mean peak VO_2_ increased by 30% following training (baseline: 452.8±134.5; 6-months: 591.8±190.3 ml/min) with increases observed in all participants in whom measurements were obtained (Figure 3A). VO_2_ data were unavailable at baseline for one participant (EX-02) due to metabolic cart equipment failure. When normalized to body weight, mean, mean peak VO_2_ increased from 14.6±4.7 to 17.3±6.9 ml/kg/min (p=0.125). Peak W (measured in all participants) increased by 34% from 20.0±11.4 to 26.8±11.4 watts (p<0.05, Figure 3B). Peak HR was unchanged (pre=177±11; 6-months=176±9 bpm). In UT DMD, peak VO_2_ declined and peak W remained unchanged over 6-months (Figure 3C and D). Between-group comparisons demonstrated significant differences in 6-month change for both peak VO_2_ (UT DMD: -29.3±60.7 versus trained DMD: +139.0±75.5 ml/min, p<0.05) and peak W (UT DMD: 0.0±4.1 versus trained DMD: +6.8±2.4 watts, p<0.05). During constant-load submaximal cycling, mean HR decreased following training (baseline: 154.2±8.3 bpm; 3-months 146.6±4.0 bpm; 6-months: 141.5±4.1 bpm p<0.01, Figure 3E). Participants also increased continuous cycling time within their prescribed HR zone from 5.8±2.0 minutes at baseline to 13.5±5.0 minutes at 3-months and 21.3±2.5 minutes at 6-months (p<0.01, Figure 3F).

**Figure 3.**
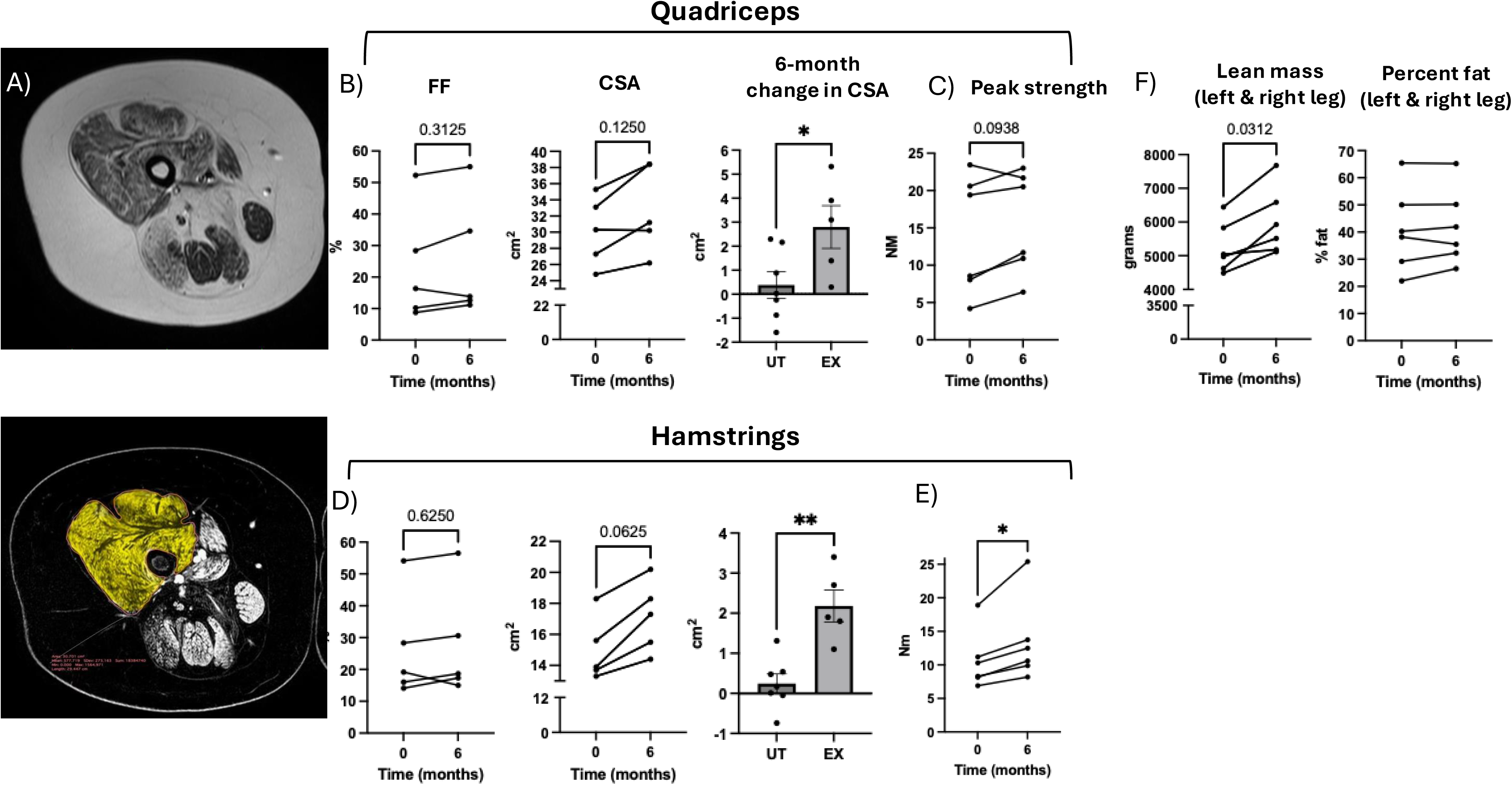
Effects of personalized exercise training on cardiorespiratory fitness in DMD. A) Peak VO_2_ before and after the 6-month intervention shown for individual participants (data available for 5 participants). For clarity, overlapping data points are noted for two participants at baseline (EX-04: 398 ml/min; EX-05: 400 ml/min) and at 6-months (EX-04: 459 ml/min; EX-05 461 ml/min). B) Peak cycling workload (W) measured in all six participants before and after training (p<0.05). C) The 6-month change in peak VO_2_ (mean ± SEM) in the exercise-trained DMD cohort compared with an untrained DMD cohort (p<0.05). D) 6-month change in peak workload (mean ± SEM) in exercise-trained and untrained DMD cohorts. E) Heart rate (HR) responses during constant load submaximal cycling shown as mean± SEM across participants, with trajectories modeling using a quadratic polynomial at baseline, 3 and 6 months. F) Time spent cycling within the prescribed HR zone before and after training. Abbreviations: HR=heart rate; UT=untrained; EX= exercise trained, VO_2_ = rate of oxygen utilization; SEM: standard error of the mean.

### Skeletal muscle and strength outcomes

Pre- and post-intervention MRI data (Figure 4) were available for 5 participants (baseline scan unavailable for EX-05). Mean FF was unchanged in the quadriceps (baseline: 23.2±18.0%; 6-months: 25.5±19.1%) and hamstrings (baseline: 26.3±16.5%; 6-months: 27.6±17.2%). CSA trended to increase in the quadriceps (baseline: 30.2±4.2 cm^2^; 6-months: 32.9±5.4 cm^2^ p=0.125) and hamstrings (baseline: 14.9±2.1 cm^2^; 6-months: 17.1±2.3 cm^2^, p=0.0625). Similar trends were detected for contractile CSA (quadriceps baseline: 23.5±7.7 cm^2^; 6-months 24.8±8.8 cm^2^; p=0.4375; hamstrings baseline: 11.1±2.9 cm^2^; 6-months: 12.5±3.6 cm^2^, p=0.0625). In the UT DMD cohort, CSA remained unchanged over 6-months. Between-group analyses demonstrated greater increases in total CSA in the trained cohort for both the quadriceps (Δ+2.8±0.9 cm^2^ versus +0.4±1.4 cm^2^, p<0.05) and hamstrings (Δ +2.1±0.89 cm^2^ versus +0.25±0.63 p<0.01). Similar results were obtained for contractile CSA in the quadriceps (Δ1.3±2.1 cm^2^ versus -0.22±0.90 cm^2^, p=0.1225) and hamstrings (Δ +1.46±0.91 cm^2^ versus +0.26±0.51 cm^2^, p=0.0519). DEXA analysis demonstrated in increase in leg lean mass in the trained cohort (Δ+768±440 grams, p<0.05) with no change in fat percentage (Δ +1.1±2.6 %, Figure 4E). Maximal KF strength increased (Δ +2.8±1.9 nm, p<0.05) whereas changes in quadriceps strength were smaller (Δ +1.7±1.9 nm, p=0.09). Ambulatory motor function measured remained stable (Δ 10-meter walk run = +1.0±1.6 seconds; Δ 6MWD = - 5.5±24 meters; Δ NSAA score = -0.8±1.8, Figure 5).

**Figure 4.**
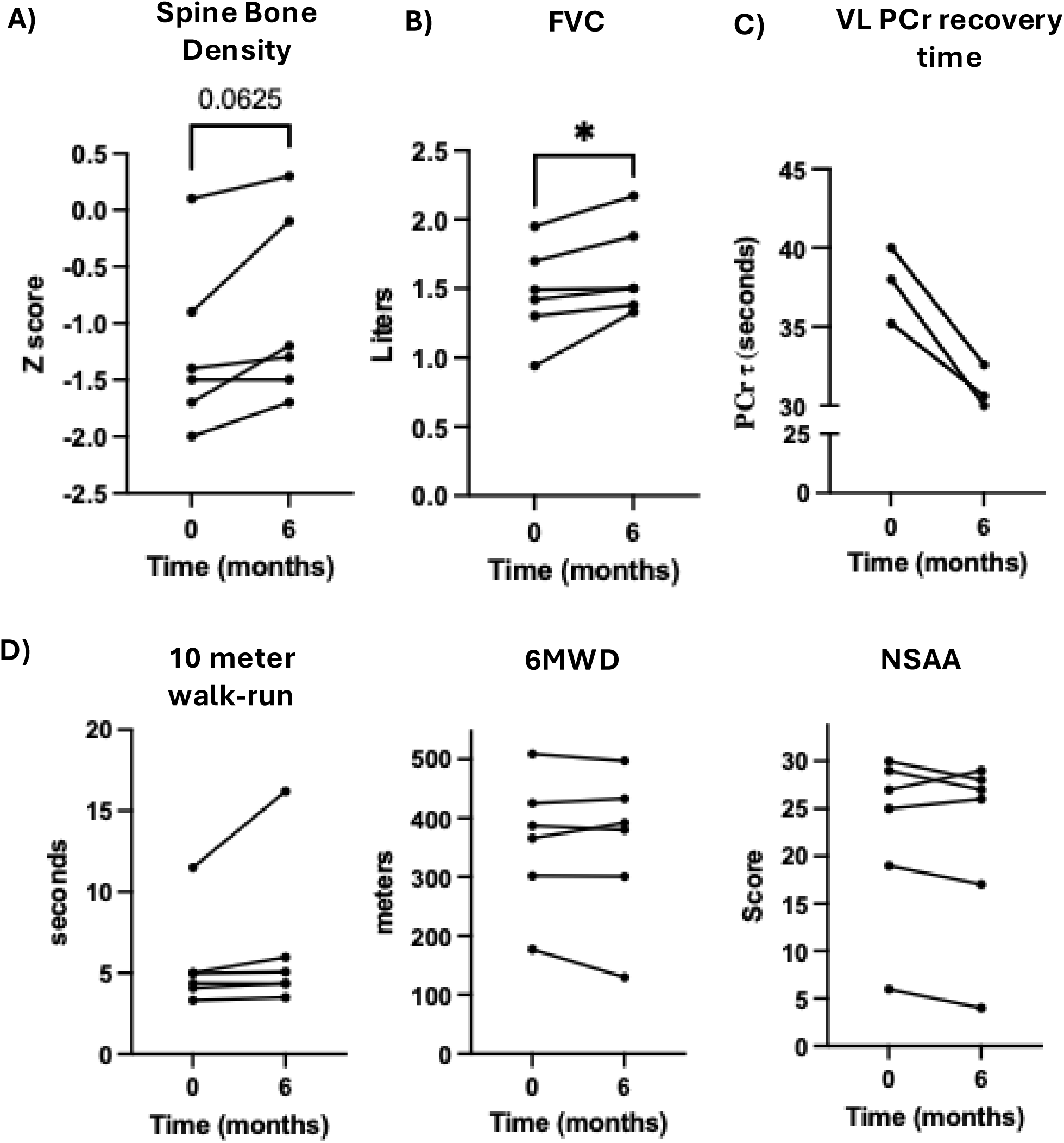
Effects of the exercise intervention on skeletal muscle outcomes in DMD. A) Representative MRI images of the upper leg illustrating regions of interest delineated in the quadriceps (yellow) and hamstring muscles for quantitative analysis. B) Individual participant changes in quadriceps muscle fat fraction and CSA at baseline and after 6 months of training. The 6-month change in quadriceps CSA is also shown for exercise-trained and untrained cohorts. C) Change in peak of knee extensors torque (p=0.0938) following the intervention. D) Hamstring muscle outcomes, including individual changes in FF and CSA at baseline and 6-months. The 6-month change in CSA is shown for exercise-trained and untrained cohorts. E) Change in peak knee flexor torque (p<0.05) following the intervention. F) DEXA-derived changes in leg lean mas and percent fat following training.

**Figure 5.**
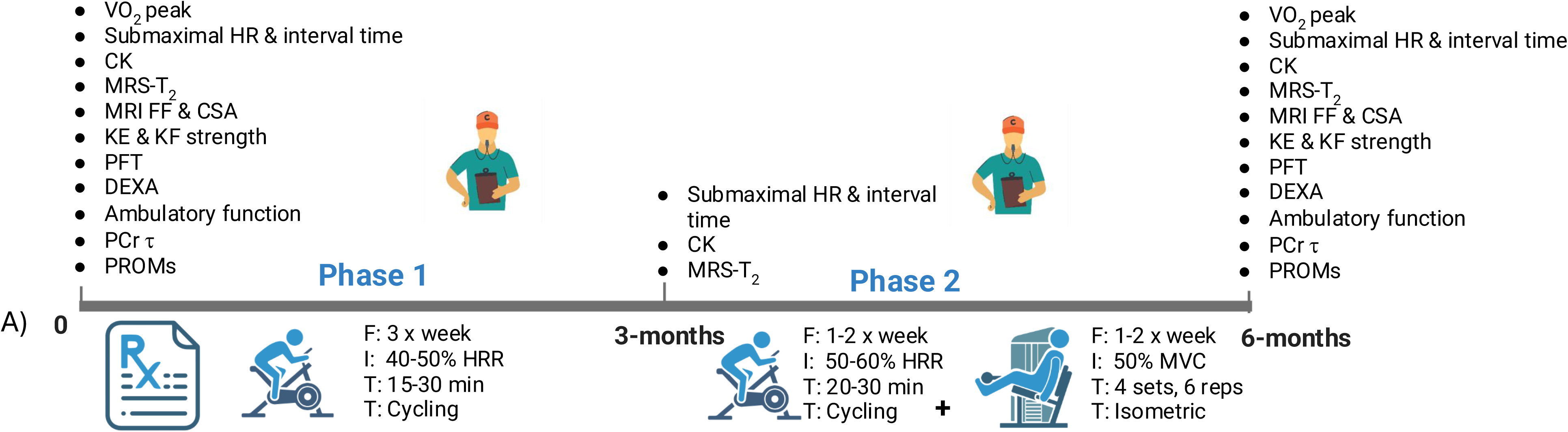
Effects of exercise training on exploratory outcomes in DMD. A) Lumbar spine bone density Z-scores are shown for individual participants at baseline and following 6-months of training. B) Forced vital capacity (FVC) measured at baseline and after completion of the intervention. C) Phosphocreatine recovery time constant assessed using ^31^P MRS in the three participants who underwent this exploratory evaluation. D) Ambulatory function outcomes, including 10 meter walk-run time, 6MWD and the North Star Ambulatory Assessment score at baseline and following the intervention. Abbreviations: FVC: forced vital capacity; MRS, magnetic resonance spectroscopy; PCr, phosphocreatine; 6MWD, 6-minute walk distance; NSAA, North Star Ambulatory Assessment.

### Pulmonary function, bone density and exploratory outcomes

Forced vital capacity increased following training (baseline: 1.46±0.34 L, 94.8±14.3% predicted; 6-months: 1.62±0.33 L, 98.3±9.23% predicted, p<0.05) with no change in FEV1 (pre: 1.31±0.32; post: 1.38±0.40). Maximal inspiratory and expiratory pressures were obtained in 4 patients revealing a trend for improvement in MIP (baseline: 49.8±14.1; 6-months 61.3±21.6 cmH_2_O, p=0.1250) while MEP was unchanged (baseline: 62.3± 21.5; 6-months: 61.0±20.8 cmH_2_O). Lumbar spine bone density Z-score improved from -1.2±0.7 to -0.91±0.8 at 6-months (p=0.0625, Figure 4E). We verified these results using height adjusted Z-score (height-for-age, HAZ) as recommended given the short stature common in DMD^51^ and observed similar improvements (pre=1.3; post=1.7, p=0.0625). ^31^PMRS was performed as an exploratory outcome in 3 participants and demonstrated faster PCr recovery kinetics (Figure 3C).

### Quality of Life

Patient-reported outcomes demonstrated improvements following training. The PedsQL-DMD total score trended to be higher after training (71.8±8.8 to 82.6±15.4, p=0.0757) while the PedsQL-MDF increased significantly (72.1±8.9 to 86.2±13.8, p<0.05). The caregiver impression of change scores averaged 5.4±0.15, corresponding to “somewhat” and “much improved”, compared with a reference value of 4 (“no change”, p<0.01). In free-text comments accompanying the impression of change, caregivers frequently noted perceived improvements in confidence, independence and participation in school-related activities.

## DISCUSSION

Exercise is considered a medicine that can prevent and treat chronic disease, with ACSM-formulated prescriptions tailored to underlying pathophysiology in a myriad of medical conditions ^30, 52–55^. However, due to susceptibility to contraction-induced injury and concerns of overuse weakness^15, 56^, exercise has not been widely adopted as a therapeutic strategy in DMD. This study sought to address this important care gap by applying a precision exercise framework, in which exercise dose and modality were individualized to mitigate risk in a cohort of ambulatory boys with DMD. Our findings support the feasibility of this approach for moderate-intensity cycling and isometric strengthening, with objective outcome measures indicating improvements in cardiorespiratory capacity and muscle strength without evidence of adverse effects on muscle pathology. Signals of benefit were also observed in bone density, pulmonary function and quality of life, supporting the pleiotropic potential of structured exercise in this population.

Prior cycling exercise studies in DMD reported safety, feasibility and potential to delay functional deterioration^16–18^, but evidence for therapeutic efficacy remains uncertain^22^. These earlier studies appropriately took a conservative approach by using motor-assisted cycling, however information on HR is lacking raising questions regarding stimulus adequacy. The prevailing view has been that muscle weakness constrains the cardiopulmonary system^57, 58^ limiting increases into a HR zone consistent with physiological adaptation. We recently demonstrated that passive, motor-assisted cycling increases HR by less than 10-bpm, whereas active cycling elicits ∼40-bpm increases, reaching target zones (40-50% HRR) consistent with ACSM recommendations for cardiorespiratory exercise benefits^28^. These findings informed the present trial, suggesting HR (gold-standard to guide exercise intensity^29, 30^) could be used to dose cycling intensity and evaluate whether individuals with DMD adapt to an appropriate exercise stimulus.

In the absence of consolidated exercise prescription guidelines for DMD, we adapted FITT parameters from deconditioned and heart failure populations^25^. These parameters are comparable to those shown to improve skeletal muscle and cardiorespiratory outcomes in metabolic myopathies with similarly reduced peak VO₂^59–61^ and align with recently proposed FITT-based recommendations for exercise in muscle disease^27, 62^. To further tailor the intervention to DMD pathophysiology and address both fatigue and weakness, we selected active cycling and isometric strengthening - modalities that minimize eccentric loading. For cardiorespiratory exercise, guidelines suggest starting with 2–3 sessions per week, 40–60% HRR for 20–30 minutes, with gradual progression of intensity and duration^25, 30^. To optimize benefit, we individualized cycling intensity using resting HR and peak HR measured during CPET rather than age-predicted estimations (e.g., peak HR = 0.7 × age^63^). Importantly, measured peak HR (177±11 bpm) was substantially lower than age-predicted values (203±1 bpm), underscoring the risk of overdosing intensity using standard equations. Inappropriately prescribed intensities can contribute to low adherence or limited efficacy^64, 65^. Isometric strengthening was dosed at 50% of MVC, an approach previously demonstrated to be safe, feasible, and effective for improving strength in DMD^20^. Adherence to these individualized FITT parameters was high (88.5%) and no-exercise related adverse events occurred. Serum CK, muscle MRS-T_2_ values and ejection fraction remained stable, suggesting no exacerbation of muscle injury with the personalized parameters: cycling 3 times per week up to 30 minutes at an average 57.5% HRR and isometric strengthening 1-2 times per week at 50% MVC. These findings support the safety and feasibility of individualizing intensity using measured HRR and %MVC rather than age-predicted estimates.

Therapeutic efficacy was assessed using peak VO_2_ and submaximal exercise HR, established markers of cardiorespiratory adaptation^25^. The impact of exercise training on these outcomes has not previously been quantified in DMD. Peak VO_2_ increased by 30%, consistent with expected improvements (10-25%) in deconditioned populations^59, 61^. This absolute increase (14.6 to 17.3 ml/kg/min) is clinically meaningful given the strong prognostic relevance of cardiorespiratory fitness (>18.0 ml/kg/min) for independence, cardiometabolic risk and survival^66–68^. Submaximal exercise HR decreased in all participants (∼13 bpm), reflecting improved exercise tolerance and myocardial efficiency^69^. Although mechanisms (i.e. increases in stroke volume, plasma volume and reductions in sympathetic drive) were not examined, the consistent improvements in both peak and submaximal cardiorespiratory indices are clinically significant in a disease characterized by progressive cardiorespiratory decline. Additionally, FVC improved and MIP trended to be higher after training suggesting benefit to pulmonary function and inspiratory muscle strength.

To date, it has not been established whether exercise can modify muscle pathology or influence disease progression in DMD. The only prior study addressing this question reported no change in muscle architecture (assessed by ultrasound) after 4-months of motor-assisted cycling^18^. We used MRI to quantify FF (a reliable and sensitive biomarker of DMD disease severity and progression^32, 40, 70^) within muscles recruited during the exercise paradigm. Baseline FF was elevated as expected in DMD (range 9-54%) and changed minimally after the 6-month intervention (quadriceps Δ+2.3%; hamstrings Δ+1.3%). These changes fall within or slightly below the expected annualized progression based on baseline FF in DMD natural history studies (∼8-10%^70^), suggesting that exercise does not accelerate fatty infiltration. Notably, in patients with Facioscapulohumeral muscular dystrophy type 1, cardiorespiratory exercise training has been shown to reduce fat infiltration to approximately half the expected annualized rate^71^.

We also evaluated the adaptive potential of dystrophic muscle to an exercise stimulus. Evidence of modest exercise-induced hypertrophy was observed with increases in quadriceps and hamstring CSA (Δ+2 to 3 cm^2^), however comparison with the untrained DMD cohort which showed no change over a similar interval, strengthens their physiological relevance. Improvements in peak isometric torque, particularly in the hamstrings (KEΔ+12%; KFΔ+26%) further support these findings. Consistent with this, average leg lean mass assessed by DEXA increased by 768 grams. ^31^P-MRS demonstrated faster PCr recovery kinetics (PCrτ) in all three of the participants who were assessed with this technique, indicating enhanced muscle oxidative capacity – an important finding given the contribution of mitochondrial dysfunction to DMD pathophysiology^72, 73^. These results align with cardiorespiratory evidence of improved exercise tolerance following training. Collectively, these findings provide preliminary *in vivo* evidence that dystrophic muscle in individuals with DMD retains capacity for favorable remodeling in response to appropriately prescribed exercise. This aligns with decades of preclinical work demonstrating beneficial effects of exercise in dystrophic muscle^74–76^, including recent data in the more severe D2.mdx mouse model showing improvements in muscle mass and mitochondrial function with voluntary wheel running^77^.

We next explored whether personalized exercise conferred benefits beyond skeletal muscle and the cardiorespiratory system by assessing its impact on bone health and patient-reported outcomes. Mechanical loading and physical activity are known to stimulate bone formation particularly in the lumbar spine of healthy children^25^ ^78, 79^. Bone health is compromised in DMD due to reduced mechanical loading associated with progressive muscle weakness^80^ and chronic corticosteroid exposure^8,81^. We observed a trend toward improved lumbar spine density Z-score following 6-months of training. Although not statistically significant, improvements in Z-score over time are infrequently reported in DMD natural history studies and may therefore be clinically meaningful. These findings suggest that the loading stimulus achieved through active cycling^82^ and isometric strengthening may be sufficient to elicit a favorable response despite long-term corticosteroids use^8^. Consistent with this broader impact, exercise is well-established to improve mood, mental well-being and quality of life^83^. In a therapeutic landscape largely focused on modifying muscle pathology, it may represent one of the few interventions capable of directly targeting psychosocial well-being. Our findings of improved caregiver and patient reported fatigue and quality of life following the 6-month exercise intervention support this notion.

We previously demonstrated that 12 weeks of isolated isometric leg strengthening in DMD increased peak torque with concurrent improvements in functional ability (time to ascend/descend 4-stairs)^20^. However, in the present study, these strength gains did not translate into measurable improvements in ambulatory and functional outcomes (10-meter walk run time, 6MWD or NSAA score). This likely reflects training specificity, as the current intervention emphasized cardiorespiratory cycling rather than ambulation. The observed increase in peak cycling power (Δ7 watts) supports this modality-specific adaptation. Importantly, these functional measures were on average maintained over 6-months, which may be considered a positive outcome in the context of a rapidly progressive disease where an annual decline of approximately 4 points in NSAA is expected after age 7^84^. In one participant (EX-06), a notable decline in ambulatory measures was observed (Figure 4C-E). Despite this, he maintained high adherence to the intervention, cycling approximately 1 mile per session 3 times per week and completing strength training close to twice weekly, demonstrating improvements across primary outcomes.

Findings from this study are limited to the small sample size of 6 ambulatory boys. This proof-of-concept study was intentionally designed to be exploratory and incorporate comprehensive, quantitative and non-invasive outcome measures to establish safety and feasibility of precision exercise in DMD prior to larger-scale investigation. A similar sample size (n=8) was used in our prior strength-training study demonstrating safety and preliminary efficacy in DMD^20^. Participants were not randomized to exercise or control groups; instead, untreated DMD controls were included as comparators to contextualize changes in primary and secondary outcomes over this time-period. The design also precluded determination of the relative contributions of each exercise modality or whether addition of strength training to cycling conferred further benefit. Mechanistic insights were beyond the scope of this study. Increases in peak VO_2_ may reflect cardiac, ventilatory and/or skeletal muscle adaptations, all of which are compromised in DMD. The observed reduction in submaximal HR may indicate improved sympathetic response to exercise and reduced myocardial workload ^69^, but the specific physiological drivers warrant further study, particularly regarding cardiac adaptations. Our findings on leg cycling and isometric strengthening benefits cannot be generalized to non-ambulatory DMD. However, observations from one participant who demonstrated substantial decline of ambulatory function yet continued to exercise using personalized parameters suggests that the precision exercise paradigm proposed here may be feasible in non-ambulatory DMD, including through upper-extremity modalities. Finally, because exercise was closely supervised, translation to the real-world is uncertain. Nonetheless, recent home-based cardiorespiratory exercise with coaching in adults with neuromuscular disease showed continued benefits on physical fitness levels over 1-year supporting the transition from therapist-supervised exercise to integration of physical activity into daily life ^85^. The inclusion of such coaching through the DMD multi-disciplinary clinical team along with exercise guidelines provided herein may facilitate translation to daily life.

In conclusion, this study establishes a framework for personalizing exercise in individuals with DMD by defining safe and feasible FITT parameters for cardiorespiratory exercise and isometric strengthening within a home-based setting. Our pilot data provide preliminary evidence that appropriately dosed exercise can elicit meaningful physiological adaptations in skeletal muscle and cardiorespiratory systems, with potential downstream benefits for bone health and quality of life. Collectively, these results add to the growing body of literature supporting the therapeutic role of exercise in clinical populations, including those with severe muscle weakness. Importantly, this work lays the foundation for future larger-scale, randomized controlled trials to delineate dose–response relationships; evaluate long-term potential to modify disease trajectory, steroid-associated side-effects and multi-systemic impairments and investigate the role of exercise as an adjunct to emerging gene and pharmacological therapeutics. Within the broader shift toward precision exercise in clinical care, an individualized approach to exercise can be envisioned in DMD, whereby exercise prescriptions are tailored to genotype, concurrent therapies, physiological status, and patient-centered goals, with ongoing monitoring of biomarkers and functional outcomes to optimize safety and maximize clinical benefit ^24^.

## Data Availability

All data produced in the present study are available upon reasonable request to the authors

## Acknowledgements

The authors thank the participants and their parents for their participation and valuable contribution to this study.

## Statements and declarations

## Ethical considerations

This study was approved by the University of Florida Institutional Review Board 1 (Project: 201901339). Written informed consent and assent were obtained for each participant. All procedures conformed to the Declaration of Helsinki.

## Consent for publication

Informed consent for publication of images for educational purposes was provided by legally authorized representatives.

## Data availability

The data supporting the findings of this study are available on request from the corresponding author.

