## Supplemental Table 1 for "Feasibility and efficacy of personalized cardiorespiratory and strength exercise in Duchenne muscular dystrophy: a pilot study"

**Supplemental Table 1:** Personalized FITT parameters for exercise prescription in DMD cohort

| | | | | | | | | | | | |
|---|---|---|---|---|---|---|---|---|---|---|---|
| | **Cycle endurance training** | | | | | | | **Isometric Strengthening** | | | |
| | **Rx** | **Phase 1** | **Phase 2** | **Total** | | | | **Rx** | **Phase 2** | | |
| **F:** | **2-3x / wk** | **Avg # per week** | **Avg # per week** | **# completed** | **Avg # per week** | **% Rx** | | **1-2x /wk** | **Avg # per week** | **# completed** | **% Rx** |
| EX-1 | | 2 | 1 | 35 | 1.5 | 58 | | | 1.5 | 19 | 79 |
| EX-2 | | 2.5 | 2 | 54 | 2.3 | 90 | | | 1.3 | 16 | 67 |
| EX-3 | | 3 | 3 | 67 | 3 | 111 | | | 1.3 | 15 | 63 |
| EX-4 | | 2.5 | <1 | 34 | 1.5 | 57 | | | 2 | 25 | 104 |
| EX-5 | | 2.5 | <1 | 32 | 1.3 | 53 | | | 3 | 36 | 150 |
| EX-6 | | 3.3 | 2 | 61 | 3 | 100 | | | 1.7 | 20 | 83 |
| **Avg± Stdev** | | **2.6±0.5** | **2±0.8** | **47±15** | **2.1±0.7** | **79±26** | | | **1.8±0.6** | **22±7.8** | **91±32** |
| | **Cycle endurance training (Phase 1 and 2)** | | | | | | | **Isometric Strengthening** | | | |
| **I:** | **40-60% HRR** | **Rest HR (Bpm)** | **Peak HR (Bpm)** | **HRR (Bpm)** | **40-60% zone (Bpm)** | **Avg HR (Bpm)** | **Avg Power (watts)** | **50% MVC** | **KE torque (Nm)** | **KF torque (Nm)** | |
| EX-1 | | 111 | 180 | 69 | 139-152 | 134±6 | 4.2 | | 8.3 | 5.8 | |
| EX-2 | | 105 | 189 | 84 | 139-155 | 155±8.8 | 6.1 | | 12.5 | 16.0 | |
| EX-3 | | 118 | 186 | 68 | 145-159 | 153±6.9 | 6.2 | | 25.0 | 28.0 | |
| EX-4 | | 102 | 173 | 71 | 130-145 | 144±14 | 5.5 | | 9.9 | 5.0 | |
| EX-5 | | 120 | 160 | 40 | 136-144 | 147±10.5 | 4.0 | | 6.9 | 10.2 | |
| EX-6 | | 110 | 173 | 63 | 135-148 | 155±6.1 | 4.4 | | 4.9 | 10.4 | |
| **Avg± Stdev** | | **111±7.0** | **177±11** | **66±14** | **137-151** | **148±8** | **5.1±1.0** | | **11.2±7.1** | **12.6±8.5** | |
| | **Cycle endurance training (Phase 1 and 2)** | | | | | | | **Isometric Strengthening** | | | |
| **T:** | **15-30 min** | **Initial interval** | **Initial duration (min)** | **Final interval time (min)** | **Final duration (min)** | **AVG per session (min)** | **AVG distance (miles)** | **4 sets,** | **Total # of reps** | | |

| | | **time (min)** | | | | | | **6 reps** | |
|---|---|---|---|---|---|---|---|---|---|
| EX-1 | | 6 | 18 | 20 | 30 | 20 | 1.2 | | 1334 |
| EX-2 | | 5 | 15 | 20 | 30 | 26 | 1.5 | | 1797 |
| EX-3 | | 7 | 18 | 30 | 30 | 27 | 1.6 | | 1265 |
| EX-4 | | 5 | 15 | 15 | 27 | 25 | 1.0 | | 1898 |
| EX-5 | | 5 | 15 | 12 | 26 | 18 | 1.3 | | 2256 |
| EX-6 | | 3 | 12 | 20 | 30 | 18 | 1.0 | | 1382 |
| **Avg± Stdev** | | **5.2±1.5** | **15.6±2.5** | **20.4±6.4** | **28.6±1.9** | **21.9±3.9** | **1.3±0.2** | | **1655±392** |
